# Age-stratified Infection Probabilities Combined with Quarantine-Modified SEIR Model in the Needs Assessments for COVID-19

**DOI:** 10.1101/2020.04.08.20057851

**Authors:** Vena Pearl Bongolan, Jose Marie Antonio Minoza, Romulo de Castro, Jesus Emmanuel Sevilleja

## Abstract

We use the age-stratified COVID-19 infection and death distributions from China (more than 44,672 infectious as of February 11, 2020) as an estimate for a study area’s infection and morbidity probabilities at each age group. We then apply these probabilities into the actual age-stratified population to predict infectious individuals and deaths at peak. Testing with different countries shows the predicted infectious skewing with the country’s median age and age stratification, as expected. We added a Q parameter to the classic SEIR compartmental model to include the effect of quarantine (Q-SEIR). The projections from the age-stratified probabilities give much lower predicted incidences of infection than the Q-SEIR model. As expected, quarantine tends to delay the peaks for both Exposed and Infectious, and to ‘flatten’ the curve or lower the predicted values for each compartment. These two estimates were used as a range to inform planning and response to the COVID-19 threat.

## 1. Introduction

From the earliest reports coming out of Wuhan, it became clear that COVID-19 is heavily biased against elderly males with pre-existing conditions. This shows the first weakness of the compartmental models like SEIR: they all assume a homogenous population. Sample runs even with a quarantine-modified SEIR model gave suspiciously high estimates for peaks of exposed and infectious. This gave the inspiration for an age-stratified probabilities of infection, which serves to give a lower bound to estimates.

## 2. Methodology

### 2.1 Estimates by Age Stratification

This is a probability game which uses data of COVID-19 patients in China **[1]**, stratified by ages. We now treat the percentages in each age group as an estimate for the corresponding probabilities of infection for people in the corresponding age group. The true probabilities are unknown, but the spread of the data from China is consistent with the virus having a bias against the elderly with pre-existing conditions. As can be expected, this will skew the Chinese distribution depending on the age distribution of the area under study, and the true distribution for the study area will be revealed as actual cases are reported. The significance of this skewing is this: since the Philippines has a median age of 25.7 **[2]**, half of our population is below 25.7, so more than half of our population will be in the ‘safer’ age groups, with lower probabilities of getting infected, and those who do get infected account for only 10.2% (see Table 1, sum of % 0-29 years old) of cases. This will be true for other countries with low median ages compared to the world average median age. We attempted to directly calculate infection probabilities per age group using Hubei Province’s estimated 2019 population of 59 million; we got non-normalized probabilities (not summing to one) with the same scatter as the estimated infection probabilities (data not shown).

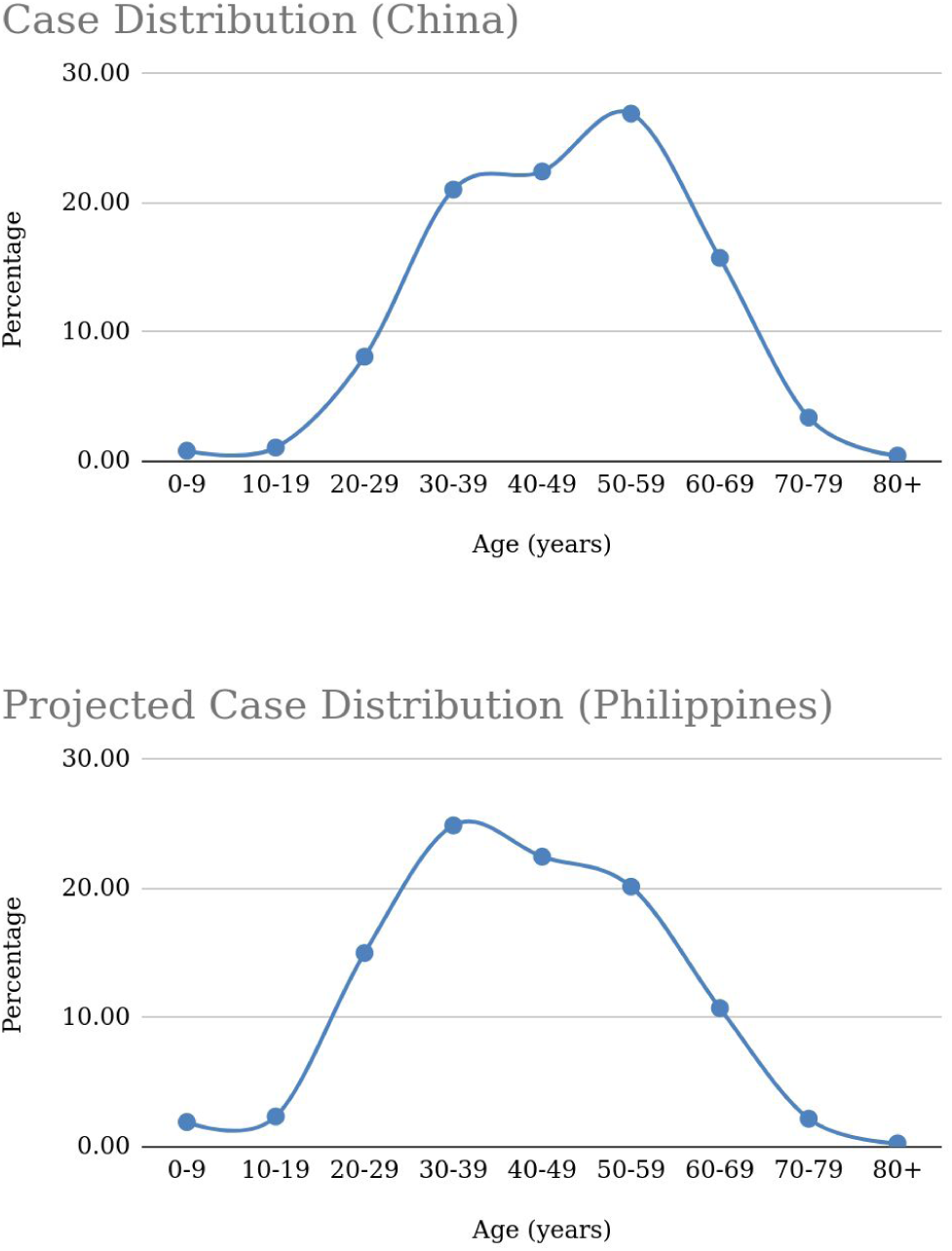

**Table 1.** Age Stratification.

| Age Groups (Years) | China (n=44,672; CFR=2.3%) |  |  | Quezon City (QC) |  |  |  |
| --- | --- | --- | --- | --- | --- | --- | --- |
|  | COVID-19 Cases (%) | Deaths (%) | CFR (%) | Pop (2020 estimate) | COVID-19 Cases | Number of Deaths | Deaths (using China's CFR) |
| 0-9 | 0.9 | 0.0 | 0.0 | 573,845 | 5,165 | 0 | 0 |
| 10-19 | 1.2 | 0.1 | 0.2 | 612,165 | 7,346 | 7 | 15 |
| 20-29 | 8.1 | 0.7 | 0.2 | 659,804 | 53,444 | 374 | 107 |
| 30-39 | 17 | 1.8 | 0.2 | 512,688 | 87,157 | 1,569 | 174 |
| 40-49 | 19.2 | 3.7 | 0.4 | 390,696 | 75,014 | 2,776 | 300 |
| 50-59 | 22.4 | 12.7 | 1.3 | 275,683 | 61,753 | 7,843 | 803 |
| 60-69 | 19.2 | 30.2 | 3.6 | 142,813 | 27,420 | 8,281 | 987 |
| 70-79 | 8.8 | 30.5 | 8.0 | 52,146 | 4,589 | 1,400 | 367 |
| ≥ 80 | 3.2 | 20.3 | 14.8 | 21,870 | 700 | 142 | 104 |
| **Total** | **100** | **100** |  | **3,241,710** | **322,587 (9.95%)** | **22,391 (6.94%)** | **2,857 (0.89%)** |

We immediately see the graph skewing to the right in the Philippines’ case, which is what we expect from a country with a younger median age (25.7, Philippines; 38.4, China, **[2]**). Using the UN World Population Prospects 2019 data **[3]**, we did a similar experiment with Japan (median age 48.6 **[2]**), and Kenya (median age 20 **[2]**).

We later found out that Martinez **[4]** did similar calculations.

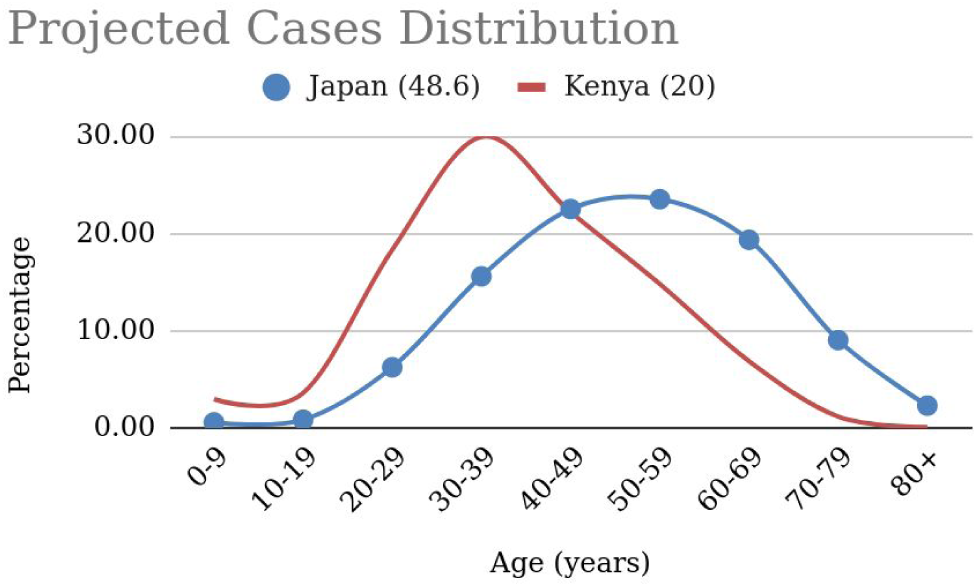

### 2.2 Estimates by a Quarantine-Modified SEIR Model

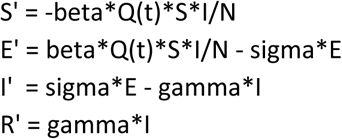

Above is the quarantine-modified SEIR. We model the quarantine as controlling the S*I interactions. A Q of one means no quarantine, and we have the original SEIR model. A Q value of 0.4 means a 60% effective quarantine. We allowed Q to vary day by day (since cases began before the quarantine), and estimated the success of the quarantine as well. Henceforth we refer to the model as Q-SEIR. Solution was via Euler method, time stepping was one day.

## 3. Results and Discussion

We use the infection probability estimates for Quezon City (QC), in the Philippines, with an age distribution as shown in Table 1 (from 2015 Census, projected to 2020 at 2% growth rate.**[6]**) and it gave an estimate of 322,586 infectious individuals (accumulated, which we equate with Q-SEIR peak), which accounts for less than 10% of the population of Quezon City. Deaths are predicted at 22,390, or 6.94% of cases, which lies between WHO morbidity estimates of 5.58% **[5]**, and the 12.47% reported in Italy **[7]**.

Preliminary reports have estimated the Philippines’ CFR to be at 4.70 (4.05 to 5.43) **[7]**. This high estimate may be explained by sampling bias, wherein severe cases may have been overrepresented because of lack of testing. Those who are infectious but are asymptomatic or who exhibit mild symptoms should also be equally represented in the testing guidelines (at the moment, they are not); not to mention those who were infectious with no symptoms and have recovered.

We tried Martinez’ calculations using CFR, which was reported at 2.3% for China **[1]**. This gave a much lower number of around 2,857 deaths, for a Quezon City CFR of 0.89%. This figure is surprisingly low, compared to the 6.94% projected using the estimated infection probability.

The delay in test reporting (est. 5-7 days **[8]**) factors in the estimation of the initial E-I-R values. In addition this delay is compounded by the incubation period and, in our opinion, moves the quarantine effect further down from the actual date of implementation (March 15^th^). We started Q-SEIR simulation on March 20, 2020 with no quarantine assumed because the steep jump in cases occurred on this date; 60% effective quarantine was set for April 2^nd^.

A portion of the worksheet is shown (Table 2), with the quarantine parameter in the second column.

**Table 2.**
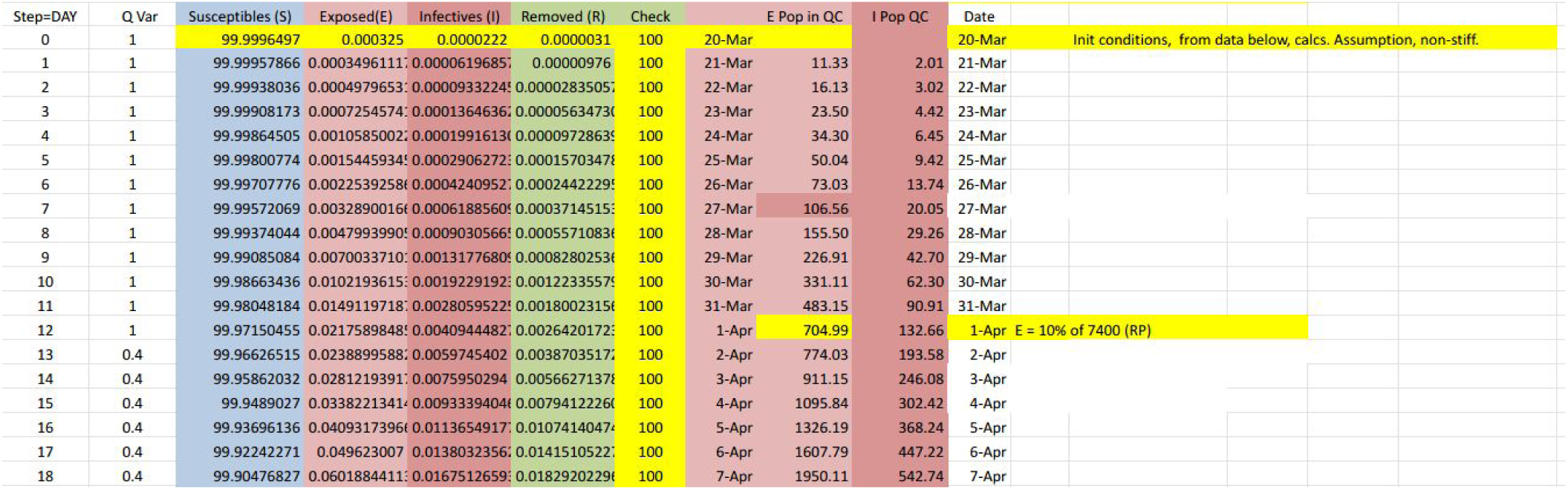
Q-SEIR.

The model was grounded to the estimated number of exposed individuals at the national level on April 1, 2020 (N=7400) **[9]**. From the nationally reported number of exposed individuals (PUI+PUM), Quezon City represents almost 10% (∼740). Q-SEIR predicted 705.

The model predicted 14.00% of the population will be infectious (I) at the peak. The two methods now give us a low and high estimate for Quezon City: Infectious individuals will peak between 9.95 (from Age Stratification) and 14.00% (from Q-SEIR) of the population, around the third week of May. This range of values serves as a guide for planners in anticipating needs for PPE’s, mass testing, hospital beds and other basic needs.

## Data Availability

Data and spreadsheets available.

## Acknowledgements

VPB thanks all her co-authors, who, up to this writing, worked pro-bono. True heroes!

## Notes

### Competing Interest Statement

The authors have declared no competing interest.

### Funding Statement

Dr. de Castro is currently supported by the Republic of the Philippines' Department of Science and Technology, Philippine Council for Health Research and Development - Balik Scientist Program.

## References

[1] The Novel Coronavirus Pneumonia Emergency Response Epidemiology Team. The Epidemiological Characteristics of an Outbreak of 2019 Novel Coronavirus Diseases (COVID-19) — China, 2020[J]. China CDC Weekly, 2020, 2(8): 113-122. URL: https://www.statnews.com/2020/03/03/who-is-getting-sick-and-how-sick-a-breakdown-of-coronavirus-risk-by-demographic-factors/

[2] Field Listing - Median age, The World Factbook, Central Intelligence Agency. URL: https://www.cia.gov/library/publications/the-world-factbook/fields/343rank.html. Last Accessed April 4, 2020

[3] United Nations, Department of Economic and Social Affairs, Population Division (2019). World Population Prospects 2019, custom data acquired via website. URL: https://population.un.org/wpp/DataQuery/ Last Accessed April 4, 2020

[4] Martinez, Ramon. Potential impact of COVID-19 in human mortality. URL: https://public.tableau.com/profile/ramon.martinez#!/vizhome/COVID-19mortalitycalculator/COVID-19mortalitycalc. Last Accessed April 6, 2020

[5] Coronavirus disease (COVID-19) Pandemic. World Health Organization. URL:https://www.who.int/emergencies/diseases/novel-coronavirus-2019, accessed April 6, 2020

[6] Quezon City. City Population. URL: https://www.citypopulation.de/php/philippines-metromanila-admin.php?adm2id=137404. Last accessed April 6, 2020.

[7] Oke, Jason and Heneghan, Carl. Global Covid-19 Case Fatality Rates. Centre for Evidence-Based Medicine Research. URL: https://www.cebm.net/covid-19/global-covid-19-case-fatality-rates/

[8] COVID-19 test results from RITM out in 5 to 7 days, but not for long, DOH says. URL:https://www.cnnphilippines.com/news/2020/3/27/COVID-19-test-results-from-RITM-out-in-5-to-7-days,-but-not-for-long,-DOH-says.html Last accessed April 1, 2020

[9] https://www.doh.gov.ph/covid-19/case-tracker Figures reported on April 1, 2020

